# Effectiveness of XBB.1.5 monovalent COVID-19 vaccine against COVID-19 mortality in Australians aged 65 years and older during August 2023 to February 2024

**DOI:** 10.1101/2024.08.12.24311895

**Authors:** Bette Liu, Anish Scaria, Sandrine Stepien, Kristine Macartney

## Abstract

**Background:** There is limited data on the effectiveness of the SARS-CoV-2 monovalent XBB.1.5 variant vaccine against COVID-19 mortality

**Methods:** We used Australian census data linked to the Australian Immunisation Register and death registrations to estimate COVID-19 booster effectiveness according to booster type and recency in adults aged 65+ years in Australia during August 2023-February 2024, a period dominated firstly by XBB-related SARS-CoV-2 Omicron subvariants and then the BA.2.86-related JN.1. Survival analysis, adjusted for age, sex, and other sociodemographic and health measures, was used to estimate vaccine effectiveness.

**Results:** We followed 4.12 million adults aged 65+ years from 1 August 2023 to 29 February 2024. By 29 February, 581146 doses of the XBB.1.5 vaccine were administered, and 1620 COVID-19-specific deaths occurred. COVID-19 mortality rates were 72/100000 person-years in people who received a COVID-19 booster >365 days earlier, and 21/100000 in those who received the XBB.1.5 booster in the last 3 months. The relative vaccine effectiveness (rVE) of XBB.1.5 booster receipt in the last 3 months against COVID-19 mortality was 74.7% (95%CI 59.9-84.1%). The rVE for those receiving other booster types in the last 3 months was 51.6% (39.3-61.4%). Booster rVE against COVID-19 mortality waned. Compared to those who received a COVID-19 booster >365 days earlier, rVE for a booster within 3-6 months earlier was 31.2% (18.9-41.6%) and for a booster received 6-12 months earlier rVE was 13.1% (1.8-23.2%). rVE estimates were similar in analyses restricted to 1 December 2023 to 29 February 2024 when the dominant Omicron subvariant was JN.1.

**Conclusions:** Recent booster vaccination with the XBB.1.5 monovalent COVID-19 vaccine is highly effective in preventing COVID-19 mortality including in the period when the JN.1 subvariant circulated, supporting recommendations for 6-monthly boosting in older adults. Evaluation of vaccination effectiveness against other health outcomes, such as COVID-19 hospitalisations and ICU admission, would help further document vaccination benefits.

## Background

In this fifth year of the COVID-19 pandemic, SARS-CoV-2 continues to evolve with immune evasion and related morbidity and mortality observed, despite widespread vaccination. In response to viral evolution, in May 2023 the WHO Technical Advisory Group on COVID-19 Vaccine Composition recommended use of a monovalent XBB.1 lineage(1). In November 2023 in Australia, the monovalent XBB.1.5 mRNA COVID-19 vaccine was registered for use in all people aged 12 years and above for primary or booster vaccination(2).

Early data from other countries has shown that the XBB.1.5 formulation is effective in preventing infection and hospitalisations from circulating SARS-CoV-2 variants(3, 4), with some studies, but not all, suggesting higher effectiveness against disease resulting from infection by XBB-lineages than the JN.1 lineage, a descendant from BA.2.86(3, 5-8). Data on the effectiveness of the XBB.1.5 vaccine against mortality from COVID-19 is more limited(6, 9). Waning of COVID-19 booster effectiveness also continues to be observed(6, 7) but how much this can be attributed to changes in viral sub-variant over time versus a decrease in individual antibody levels and immune responsiveness is difficult to disentangle due to these factors being temporally related.

Australia’s COVID-19 vaccine program has focussed on prevention of serious illness and death. Six monthly COVID-19 boosters are recommended for those aged 75 years and older. Guidelines also recommend 6-monthly boosters can be considered among those aged 65-74 years, and in adults with significant co-morbidities(10). XBB 1.5 vaccines produced by Pfizer and Moderna were recommended for use in Australia from the 20 November 2023(2). To add to the limited evidence regarding XBB.1.5 COVID-19 vaccine effectiveness against mortality, we sought to examine the effectiveness of COVID-19 boosters, recency of boosting, and booster type against COVID-19 mortality in adults aged 65 years and older in Australia during August 2023 to February 2024, a period firstly dominated by the XBB-related SARS-CoV-2 Omicron subvariants including EG.5, and then the BA.2.86-related JN.1 subvariant(5).

## Methods

The data and methods we use to assess COVID-19 vaccine effectiveness on mortality have been described previously(11). Briefly, a whole of population cohort was created using the Australian 2021 Census data linked to the Australian Immunisation Register (AIR) and national death registry through the Person Level Integrated Data Asset (PLIDA) managed by the Australian Bureau of Statistics(12). Census data include information on inhabitants of over 96% of all Australian dwellings. The AIR records all vaccines administered in Australia. Reporting of COVID-19 vaccines is mandated. Death registrations include records of all deaths registered in Australia including the contributing causes of death coded according to the WHO International Classifications of Diseases version 10 (ICD-10). Other linked data are also available to ascertain health service and medication use. For this analysis we included all individuals recorded in the Census who were aged 65 years and older and who had not migrated or died by study commencement on the 1 August 2023.

The study outcome was COVID-19 mortality defined as a death registration where the underlying cause of death was recorded as ICD-10 code U07.1 or U07.2. Individuals were classified according their COVID-19 vaccine receipt. By 2024 most older people in Australia were vaccinated, and those without significant immunocompromising conditions could have received up to 7 doses of a COVID-19 vaccine, including 2 for their primary course and up to 5 boosters. We therefore classified and compared vaccination status according to recency of receipt of a booster dose (>365 days [reference group], 181-365 days, 91-180 days, 8-90 days). Given that the XBB.1.5 vaccine was only available from November 2023, for those who had received a booster in the last 8-90 days, we also classified whether this was the XBB.1.5 monovalent vaccine or another COVID-19 booster type.

Individuals were followed from the 1 August 2023 to 29 February 2024 for COVID-19 specific mortality analysed using Cox proportional hazards models. Follow-up time was censored at date of death, receipt of an 8^th^ COVID-19 vaccine dose or 29 February 2024 whichever came first. In analyses, vaccination status was considered a time-varying covariate. Analyses were adjusted for age, sex, jurisdiction of residence, household income, number of comorbidities (assessed using an index based on pharmaceutical prescribing)(13), number of general practice visits (as a measure of health service access) and receipt of an influenza vaccine in the year prior. Adjusted hazard ratios (aHR) and 95% confidence intervals were estimated and vaccine effectiveness calculated using the formula (1-aHR) * 100%. Unadjusted Quasi-Poisson regression was used to compute crude rates of COVID-19 death.

Analyses were conducted as part of monitoring the Australian Government’s COVID-19 vaccine program and the Sydney Children’s Hospital Human Research Ethics Committee granted this surveillance work exemption from ethical review. All data used were deidentified and all results where counts or rates are provided are perturbed according to the Australian Bureau of Statistics methods to prevent disclosure of small numbers and potential re-identification.

## Results

There were 4.119 million people aged 65 years and older on 1 August 2023 who were included in the analyses. Of the included population, 45.8% (N=1.886 million) were aged 75 years and older, 53.7% (N=2.210 million) were women, 59.8% (N=2.465 million) had a household income of <1000 Australian dollars a week, 24.8% (N=1.022 million) had 6 or more comorbidities, 31.0% (N=1.277 million) had 13 or more visits to a GP in the last year and 77.4% (N=3.188 million) had an influenza vaccine reported in the previous year.

Figure 1 shows the proportion of the population according to the number of vaccine doses received on 1 August 2023 and on 29 February 2024 corresponding to the beginning and end of study follow-up. The number and proportion of individuals aged 65+ years who were unvaccinated, or had received one or two COVID-19 vaccine doses remained stable across this period at 11% of the population (n=459,049 on 1 August 2023). Among individuals who had received at least one booster (dose 3 or more) the number of boosters received increased with 0.9% receiving a 6^th^ vaccine dose in August 2023 and growing to 18.8% by 29 February 2024.

**Figure 1:**
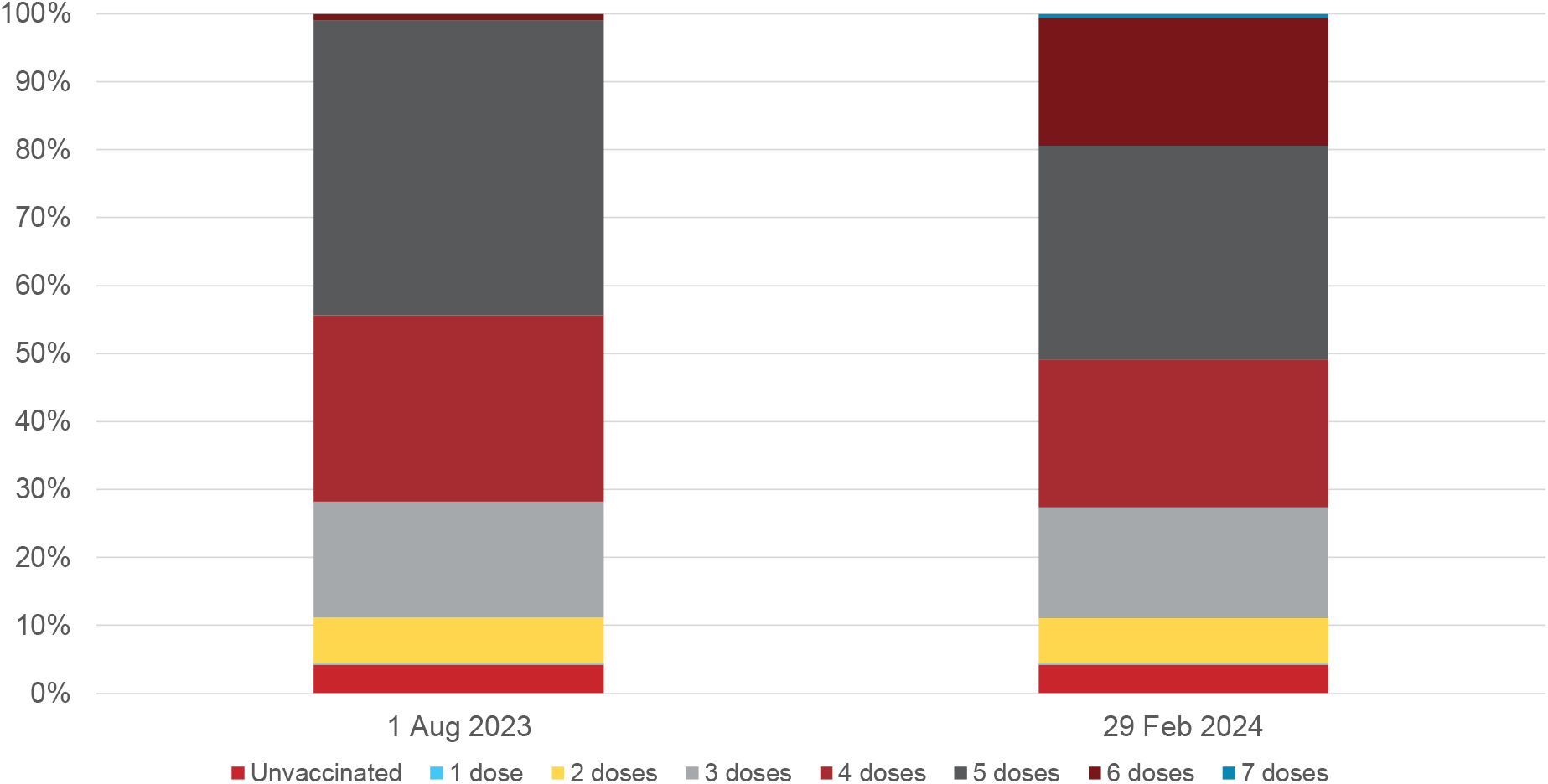
Proportion of population receiving COVID-19 vaccine doses in August 2023 and February 2024 among adults aged 65+ years

Figure 2 shows the type of booster received according to dose number for those who had a least one booster by 29 February 2024. There were 3.66 million people who had received at least 3 COVID-19 vaccines doses, 2.99 million at least 4 doses, 2.09 million at least 5 doses, 798,635 at least 6 doses and 23,996 had received 7 doses. More than half of all the boosters given for dose 6 or dose 7 were the monovalent XBB.1.5 mRNA formulation whilst dose 5 was mostly the bivalent ancestral and BA4/5 mRNA vaccine and dose 3 and 4 the original ancestral mRNA vaccine.

**Figure 2:**
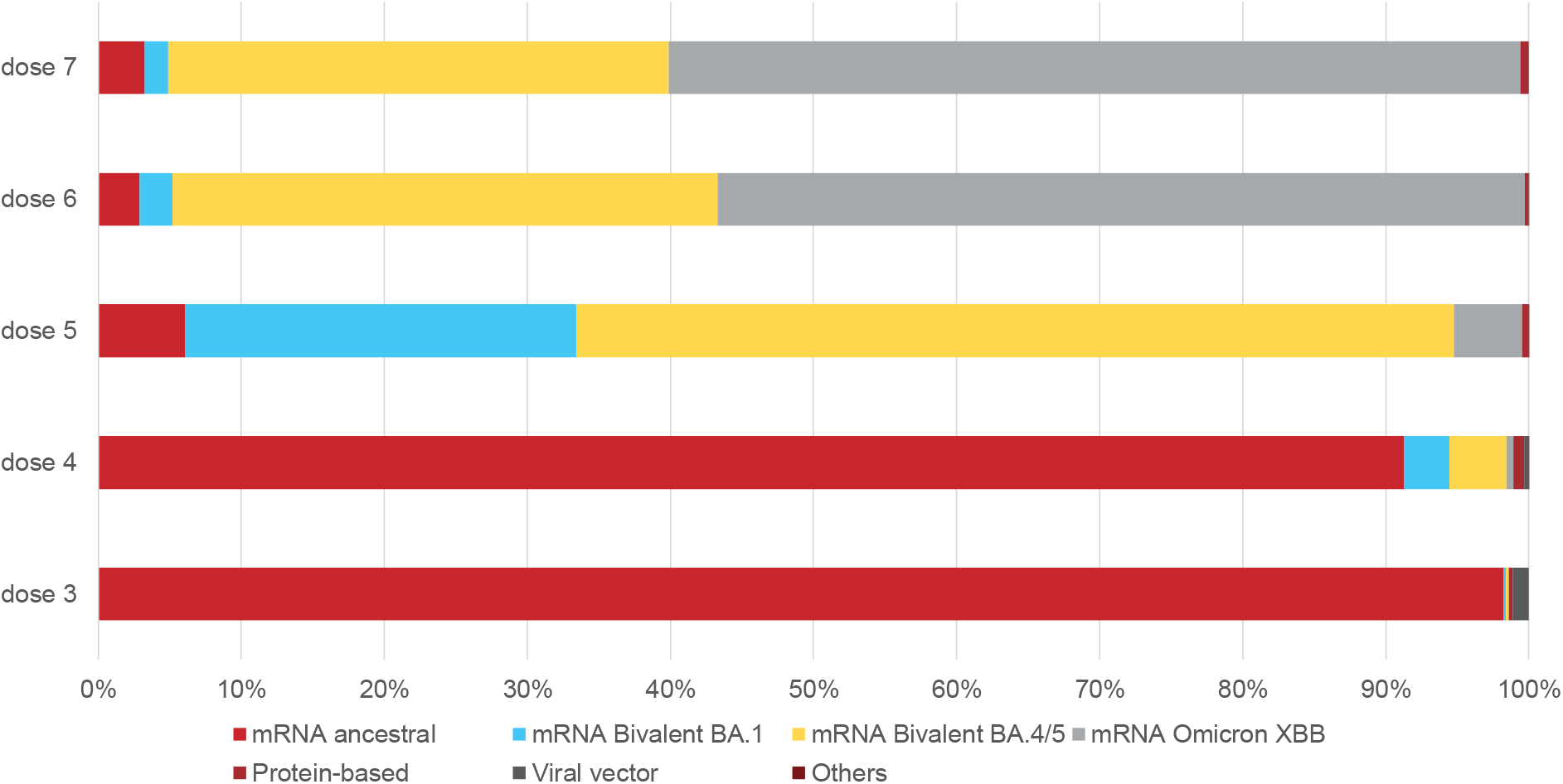
Booster vaccine type according to vaccine dose in adults 65+ years at 29 February 2024

Figure 3 shows COVID-19 vaccine effectiveness against COVID-19 mortality from 1 August to 29 February 2024. Over 2.37 million person-years of follow-up, 1620 COVID-19 deaths occurred. COVID-19 mortality rates were lower amongst those who had received a booster more recently with mortality 72/100,000 person-years in people who received a COVID-19 booster more than 365 days earlier and 21/100,000 in those who received the XBB.1.5 booster in the last 3 months. Compared to individuals who received a COVID-19 booster at least one year earlier, the relative vaccine effectiveness (rVE) of the XBB.1.5 booster (received in the last 3 months) was 74.7% (95%CI 59.9-84.1). This rVE was higher than that estimated among those receiving other booster types in the last 3 months, rVE 51.6% (95%CI 39.3-61.4%). However the average of the median time since receipt of vaccine among those who received an XBB.1.5 booster compared to those who received other booster vaccine types in the last 3 months was 32.2 days less (see Figure 4). Relative VE also waned with increasing time since vaccine receipt with rVE 31.2% (18.9-41.6%) for a booster within 3-6 months and 13.1% (95%CI 1.8-23.2%) for a booster received 6-12 months earlier.

**Figure 3:**
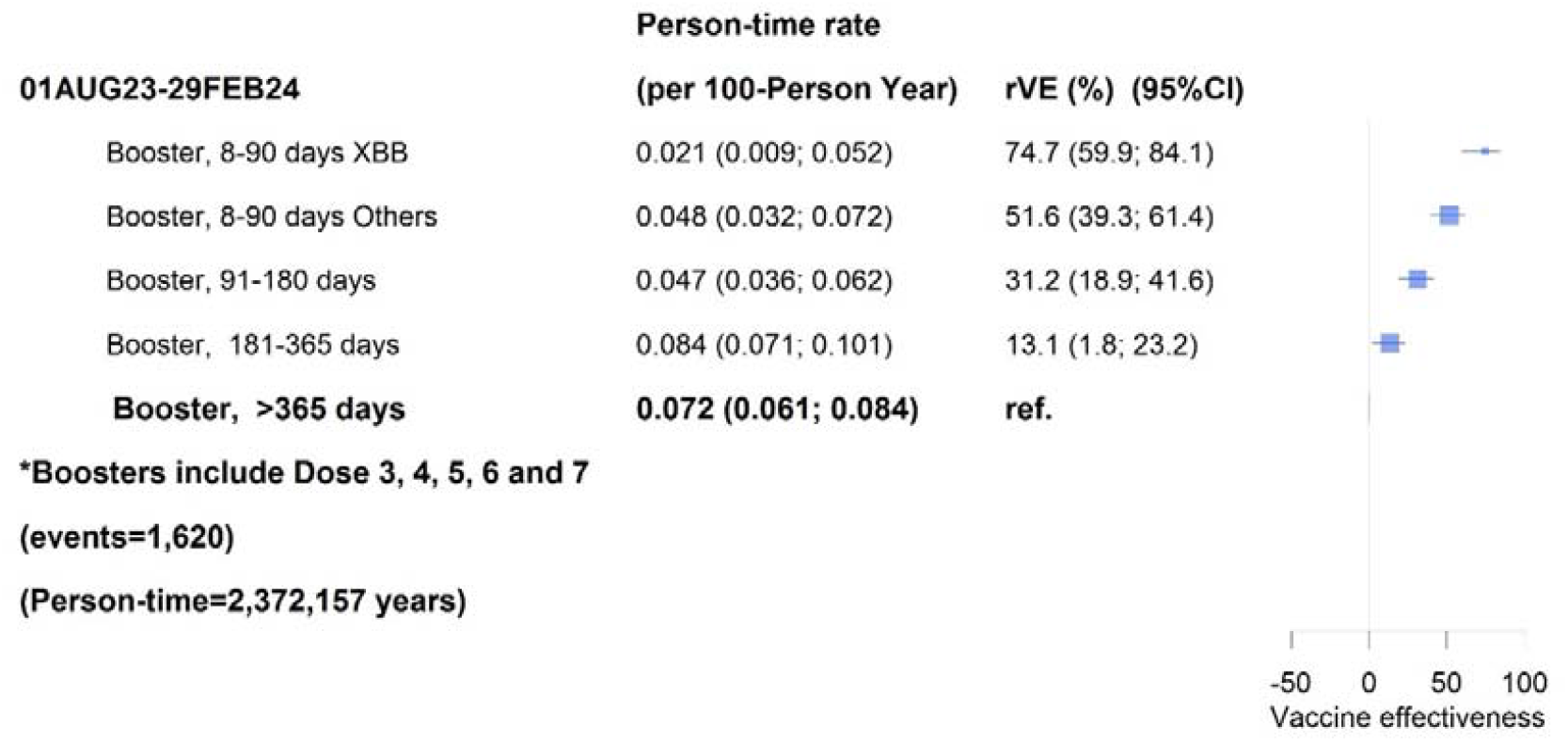
Effectiveness of COVID-19 vaccine boosters against COVID-19 mortality according to time since booster receipt in adults aged 65+ years from 1 August 2023 to 29 February 2024

**Figure 4:**
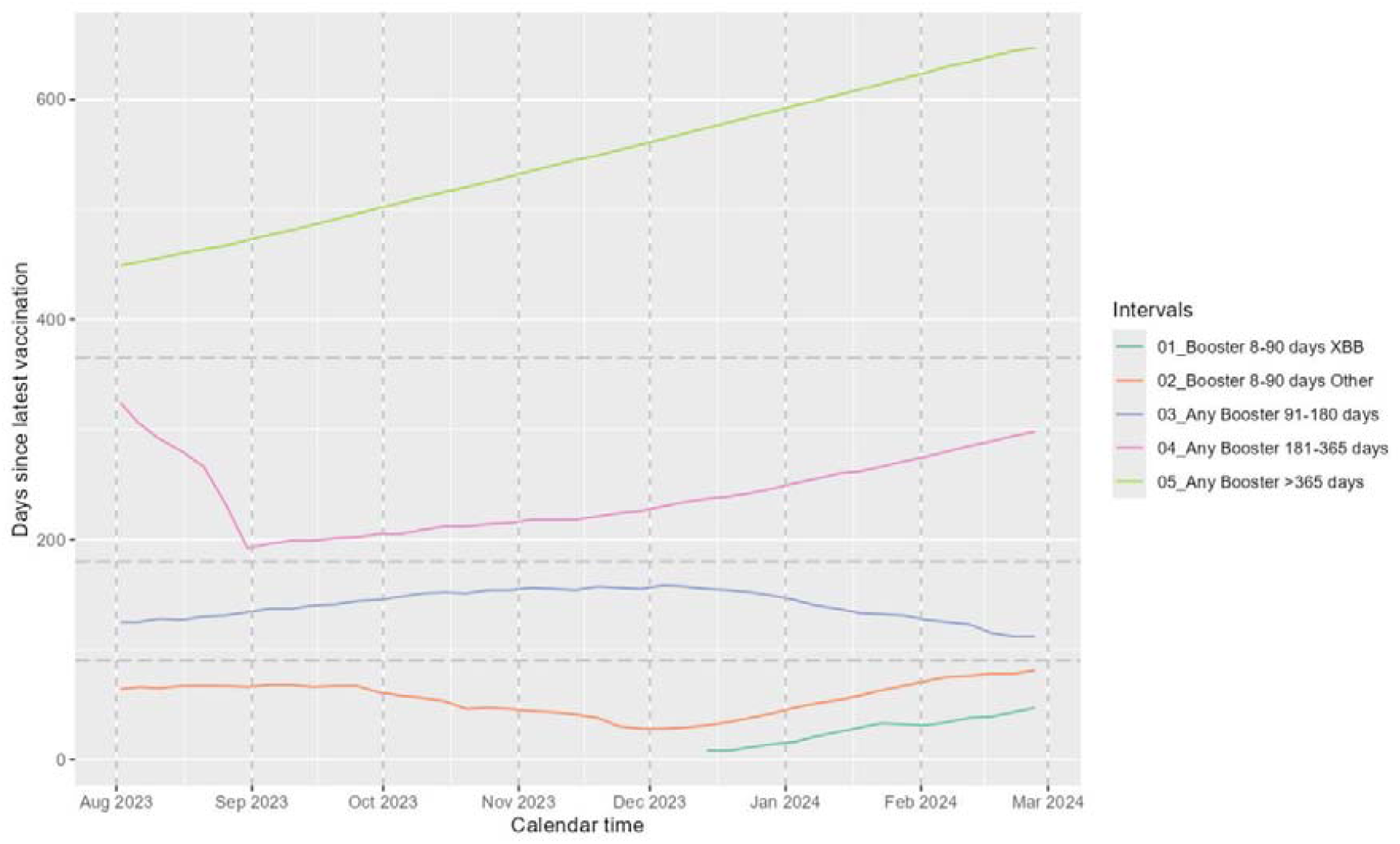
Median time since receipt of booster vaccine within categories of time since booster receipt, study time and vaccine type adults aged 65 years and over from 1 August 2023 to 29 February 2024

These finding did not materially differ when analyses were restricted to those aged 75 years and older who under Australian guidelines, were all recommended to receive 6-monthly boosters. However COVID-19 mortality rates were substantially higher in this older age group at 153/100,000 person-years in those who received a booster more than a year ago compared to 40/100,000 in those who received the XBB.1.5 booster in the last 3 months highlighting the higher population benefit of vaccination in older adults. We also found our estimates of rVE were similar to the main analysis when we restricted follow-up to the 3 month period from 1 December 2023 to 29 February 2024 when JN.1 subvariants dominated(5) (see Figure 5).

**Figure 5:**
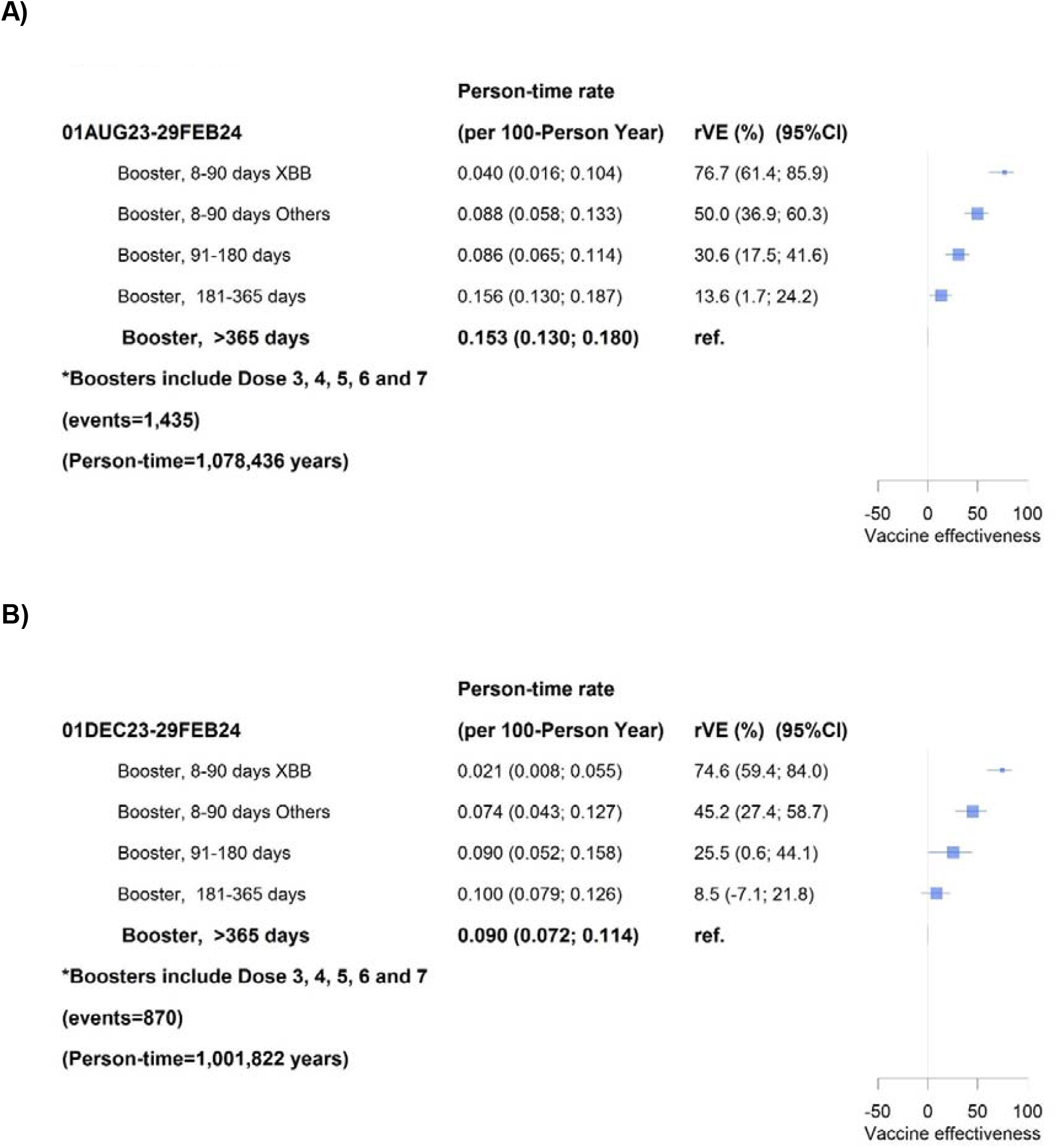
Effectiveness of COVID-19 vaccine boosters against COVID-19 mortality according to time since booster receipt in A) adults aged 75+ years from 1 August 2023 to 29 February 2024 and B) in adults aged 65+ years from 1 December to 29 February 2029

## Discussion

Monitoring the effectiveness of new COVID-19 vaccines such as the XBB.1.5 monovalent vaccine is important particularly with continuing evolution of SARS-CoV-2. We found that among adults aged 65 years and older, recent boosting with the XBB.1.5 vaccine reduced the likelihood of COVID-19 death by 75% compared to people for whom it was more than a year since their last booster receipt. We also found benefits for recent boosting with other COVID-19 vaccine types but this appeared to be lower than for the monovalent XBB.1.5. We found that the estimate of relative vaccine effectiveness against mortality was similar among adults aged 75 years and older but the absolute benefits in terms of reduction in mortality was substantially higher. We also found similar relative vaccine effectiveness during the period December 2023 to February 2024 when the dominant circulating sub-variant was JN.1 (of the BA.2.86 lineage).

Recent studies from the US, UK and Europe have consistently demonstrated that XBB.1.5 variant vaccines are effective in the short term against SARS-CoV-2 infection and hospitalisation(3, 4, 6-9, 14). There are fewer studies examining COVID-19 mortality as an outcome(8) but a large Nordic analysis suggested vaccine effectiveness is likely higher for death compared to hospitalisation. Studies are less in agreement when examining waning of the XBB.1.5 vaccine effectiveness against COVID-19 hospitalisation and effectiveness against specific SARS-CoV-2 subvariants. Some studies suggest significant waning of XBB.1.5 effectiveness in the 3 months after receipt(7), but others little waning over the same time(8). Also, effectiveness of the XBB.1.5 vaccine against infection and hospitalisation has been reported to be lower against the JN.1 sub-lineage than XBB lineages(3, 6) but this is not consistently observed in all studies(8). However, characteristics of vaccine effectiveness such as waning are much harder to estimate because changes in the circulating viral variant will coincide with increasing time since a variant-specific vaccine is introduced into a vaccine program and population.

While direct comparisons are difficult to make due to variations in the comparator groups, our estimate of the relative effectiveness of the XBB.1.5 vaccine against COVID-19 mortality is comparable to that from many other studies. For a similar average time since vaccine receipt, other studies report vaccine effectiveness against hospitalisation ranges from 55-67%(6, 8, 9, 14) and COVID-19 death 67-78%(8, 9). We did not find the relative effectiveness of the XBB.1.5 vaccine against COVID-19 death differed when only considering the period during which the JN.1 sub-variant dominated circulation (December 2023 to February 2024) although in Australia the XBB.1.5 vaccine was primarily only available during this period. Our estimates of the relative effectiveness of a booster given within 8-90 days, suggested that other booster types (which would mostly be the bivalent mRNA vaccines) were lower than XBB.1.5 (rVE 51.6% vs 74.7% (see Figure 3)). This difference has been suggested in other reports(7) but a statistically significant difference has not been demonstrated. When we examined the median number of days since receipt of vaccine among those who received vaccine 8-90 days earlier, we did find it was on average 32 days less for those who received the XBB.1.5 booster compared to other booster types. Given that we and others(7) have observed waning of mRNA COVID-19 vaccine effectiveness, it is possible the difference we observed between vaccine types could still be explained by waning over time rather actual differences in effectiveness of vaccine formulation.

Our study strengths include using large national datasets independently collected such as Census and migration data and the Australian Immunisation Register where mandatory reporting of COVID-19 vaccines has been in place since March 2021 when the COVID-19 vaccine program commenced. Our outcome of death registrations based on death certification also avoids some, but not all, biases that occur with the outcome of hospitalisation for COVID-19 where ascertainment has changed over time with changes in testing, reporting and disease severity. Limitations of observational studies of COVID-19 vaccine effectiveness are well described and include unmeasured and residual confounding from characteristics such as healthy vaccinee effects, the propensity to be more frequently boosted due to existing co-morbidities, health service access and other sociodemographic characteristics. We minimised these effects by adjusting for multiple potential confounders that were accessible in the datasets and making relative vaccine effectiveness estimates.

Overall our findings show that the XBB.1.5 variant COVID-19 vaccine is highly effective as a booster to prevent COVID-19 mortality in adults aged 65 years and older. We continued to observe waning effectiveness of COVID-19 boosters in older adults, with effectiveness more than 6 months since a booster similar to that among those who only received a booster over a year ago. Based on these findings and other data, vaccine use under current Australian recommendations to re-boost high risk populations at 6 monthly intervals will significantly reduce COVID-19 mortality and promotion of the benefits of COVID-19 vaccines particularly to those aged 75 year and above will save lives. With new recommendations regarding future COVID-19 vaccine variant formulations(15), timely assessments of COVID-19 vaccine effectiveness against severe outcomes such as mortality but also hospitalisation and intensive care admission is important to ensure our vaccine programs are based on the best evidence.

## Data Availability

All data used for this study are available upon reasonable request to the data custodians of the PLIDA resource.

## Acknowledgements

We acknowledge staff and funding from the Health Economics Research Division in the Australian Government Department of Health and Aged Care and the NHMRC Medicines Intelligence Centre for Research Excellence.

